# Induction, decay, and determinants of functional antibodies following vaccination with the RTS,S malaria vaccine in young children

**DOI:** 10.1101/2022.05.27.22275653

**Authors:** Gaoqian Feng, Liriye Kurtovic, Paul A. Agius, Elizabeth Aitken, Jahit Sacarlal, Bruce Wines, P. Mark Hogarth, Stephen Rogerson, Freya J. I. Fowkes, Carlota Dobaño, James G. Beeson

## Abstract

**BACKGROUND:** RTS,S is the first malaria vaccine recommended for implementation among young children at risk. However, vaccine efficacy is modest and short-lived. Antibodies play the major role in vaccine-induced immunity, but knowledge on the induction, decay, and determinants of antibody function is limited, especially among children. Antibodies that promote opsonic phagocytosis and other cellular functions appear to be important contributors to RTS,S immunity.

**METHODS:** We studied a phase IIb trial of RTS,S/AS02 conducted young children in malaria endemic regions of Mozambique. We evaluated the induction of antibodies targeting the circumsporozoite protein (CSP, vaccine antigen) that interact with Fcγ-receptors (FcRγs) and promote phagocytosis (neutrophils, monocytes, THP-1 cells), antibody-dependent respiratory burst (ADRB) by neutrophils, and natural killer (NK) cell activity, as well as the temporal kinetics of responses over 5 years of follow-up (ClinicalTrials.gov registry number NCT00197041).

**RESULTS:** RTS,S vaccination induced CSP-specific IgG with FcγRIIa and FcγRIII binding activity and promoted phagocytosis by neutrophils, THP-1 monocytes, and primary human monocytes, neutrophil ADRB activity, and NK cell activation. Responses were highly heterogenous among children, and the magnitude of neutrophil phagocytosis by antibodies was relatively modest, which may reflect modest vaccine efficacy. Induction of functional antibodies was lower among children with higher malaria exposure. Functional antibodies largely declined within a year post-vaccination, consistent with the decline in vaccine efficacy over that time, and decay rates varied for different antibody parameters. Biostatistical modelling suggested IgG1 and IgG3 contribute in promoting FcγR binding and phagocytosis, and IgG targeting the NANP-repeat and C-terminal regions CSP were similarly important for functional activities.

**CONCLUSIONS:** Results provide new insights to understand the modest and time-limited efficacy of RTS,S in children, and the induction of antibody functional activities. Improving the induction and maintenance of antibodies that promote phagocytosis and cellular functions, and combating the negative effect of malaria exposure on vaccine responses are potential strategies for improving RTS,S efficacy and longevity.

## BACKGROUND

Developing a highly protective malaria vaccine is a global priority given there are >240 million clinical cases of malaria and >600,000 deaths annually, which occur predominantly in young children and in endemic regions of sub-Saharan Africa [1]. Most malaria cases and deaths are attributed to infection with *Plasmodium falciparum*. The World Health Organization (WHO) and funding partners have set a goal to develop a vaccine with ≥75% efficacy (over 2 years) by 2030, but this has proven challenging to achieve.

Currently, RTS,S is the only malaria vaccine that has completed phase III clinical trials in African children, and in October 2021 the WHO recommended widespread use of RTS,S among children in sub-Saharan Africa and other regions with moderate to high *P. falciparum* malaria transmission [2, 3]. However, RTS,S demonstrated only modest efficacy with relatively short longevity. In young children aged 5-17 months, RTS,S vaccine efficacy against clinical malaria was ∼50% over 18 months of follow-up [4, 5]. Efficacy waned quickly such that there was little or no significant efficacy after 18 months [6]. When a booster dose was given at 18 months, vaccine efficacy over 4 years was 36% [7-9]. Notably, other vaccine candidates tested in clinical trials have either failed to confer significant protection in target populations of malaria-exposed individuals or have not yet demonstrated higher efficacy or greater durability than RTS,S in young children that is reproducible in multiple trials [10, 11]. Detailed knowledge is needed of immune mechanisms and kinetics of vaccine responses to address the two key limitations of RTS,S and other vaccines in clinical trials: modest efficacy and short longevity of protection. Understanding factors that impact the induction of immune responses, and determining the kinetics of antibody induction and decay could inform the refinement of RTS,S or the development of new vaccines with greater efficacy and durability [10, 12].

The RTS,S vaccine is based on the circumsporozoite protein (CSP), a major surface-expressed antigen on the sporozoite stage of *P. falciparum* [13]. The vaccine construct is composed of the central repeat region (NANP-repeats) and C-terminal (CT) region of CSP fused with the hepatitis B surface antigen and expressed as a virus-like particle [14]. Antibodies to the CSP vaccine antigen are the main mediator of protection induced by the RTS,S vaccine, although CD4+ and CD8+ T cells may also contribute to the protective response [10]. The NANP-repeat [14] and CT regions [15] are both targets of RTS,S vaccine-induced antibodies in children [14, 16, 17], and high levels of immunoglobulin G (IgG) to these domains have been broadly associated with protection against clinical malaria in RTS,S vaccine trials [13]. However, the mechanisms of action of these antibodies are not fully understood, especially among children. Furthermore, there is very limited data on the longevity of functional antibodies after vaccination. This lack of knowledge impedes progress to improve on RTS,S and constrains the development of more efficacious second-generation vaccines.

An immunologic mechanism that is being increasingly recognised in immunity against pathogens is the ability of IgG to interact with Fcγ-receptors (FcγR) expressed on various cell types leading to opsonic phagocytosis and other effector mechanisms [18-22]. We recently, identified that neutrophils were the major cell type to mediate opsonic phagocytosis of *P. falciparum* sporozoites in blood, whereas monocytes contributed a minor role [23]. Opsonic phagocytosis of sporozoites by the THP-1 monocyte cell line has also been demonstrated [19, 23]. Neutrophils express FcγRIIa, IIIa and IIIb on their surface, but may upregulate FcγRI expression in certain inflammatory settings [24]. Resting-stage monocytes predominantly express FcγRI and FcγRIIa, with a small subset expressing FcγRIIIa [25]. IgG interactions with FcγRIIa and FcγRIII are required for maximum neutrophil phagocytic activity with little requirement for FcγRI. IgG1 and IgG3 are the most potent IgG subclasses for interactions with FcγRs, while less functional activity occurs with IgG2, and little or none for IgG4. RTS,S-induced IgG1 and IgG3 responses to the NANP-repeat and CT regions were associated with protection in children [26], supporting the potential for FcγR-mediated effector mechanisms. In studies of RTS,S vaccination in malaria-naïve adults who were then challenged with a controlled experimental infection (using the homologous *P. falciparum* strain), those with higher antibody-mediated phagocytosis by neutrophils and FcγRIIa and FcγRIII binding by IgG were associated with protection from infection [18, 27]. Induction of these responses by RTS,S in young children in malaria-endemic settings is currently unknown.

Here, we investigated the induction and longevity of IgG that can interact with FcγRs and promote phagocytosis in a phase IIb trial of RTS,S/AS02 among Mozambican children residing in the Manhiça and Ilha Josina districts. The participants received the primary 3 dose regimen without any subsequent booster doses. The two cohorts represent young children exposed to different malaria transmission intensities. We aimed to quantify the ability of RTS,S vaccine-induced IgG in children to mediate interactions with activating receptors FcγRIIa and FcγRIII and promote phagocytosis, and to identify the regions of CSP that are targeted by the functional antibodies. Furthermore, we investigated how age and malaria exposure influenced the induction of functional antibodies. We applied bio-statistical models to determine the rate of decay of functional antibodies over 5 years of follow-up, and to estimate the contributions of different IgG subclasses and antibodies to different CSP regions to antibody functional activity. Studying functional responses in this phase IIb trial had the advantage of greater sampling and extended follow-up compared to the phase III trial, and no booster vaccination was administered.

## METHODS

### Study population

Serum samples collected from a previously published RTS,S/AS02_A_ phase IIb clinical trial (ClinicalTrials.gov registry number NCT00197041) were analysed in this study. This clinical trial was conducted at two study sites in Mozambique during April 2003 and May 2004 [28], whereby the Manhiça study site had low to moderate malaria transmission and the Ilha Josina study site had high malaria transmission. Briefly, children aged 1-4 years were randomised and administered 3 doses of RTS,S/AS02 (at months 0, 1, and 2) or a non-malaria comparator vaccine (children <24 months received the pneumococcal conjugate vaccine and one dose of Haemophilus influenzae type B vaccine, children >24 months received the hepatitis B vaccine). We tested serum samples from a random selection of children from the Manhiça (RTS,S, n=50; comparator, n=25) and Ilha Josina study sites (RTS,S, n=49; comparator, n=24) collected at baseline (M0) and after vaccination (M3). However, there was only sufficient sample remaining for 15 children at the M3 time point for testing ADRB and 22 children for testing in phagocytosis with primary monocytes. Sample sizes for all assays and analyses are indicated in the figure legends and the Additional file. Additional samples were collected at later time points (M8.5, M21, M33, M45 and M63) from 30 children in the Manhiça study site and used to quantify responses over time from M0 through to M63. Malaria incidence during follow-up has been previously reported [7, 28]. Over the first 6 months after the 3rd vaccine dose, 38% of children had a recorded event of fever with *P. falciparum* parasitemia; and 18% between M8.5 and M21 time-points [7].

### Research ethics

Ethics approval for this study was obtained from Mozambican National Health and Bioethics Committee; Hospital Clınic of Barcelona Ethics Committee; PATH Research Ethics Committee; Alfred Health Human Research and Ethics Committee (protocol number 174/18). Parents/guardians of participants provided written informed consent.

### Antigens

Details of the antigens used in this study were described previously [17, 29]. Briefly, CSP were expressed in *Escherichia coli* based on the 3D7 allele *P. falciparum* with the signal peptide and glycosylphosphatidylinositol sequences excluded [29][30] (produced by Gennova Biopharmaceuticals, India; provided by PATH Malaria Vaccine Initiative, USA). The C-terminal region of CSP was based on the 3D7 allele *P. falciparum* CSP and expressed in HEK293 cells [17]. A synthetic peptide of (NANP) ×15 were supplied by Life Tein (USA) to represent the central repeat region of CSP [31].

### Fcγ receptor binding assay

The FcγR binding assay was conducted using a previously published method [32]. Briefly, the dimer of the H131 allele FcγRIIa or V158 allele FcγRIII ectodomains were expressed in HEK293 cells and purified using a size-exclusion column, as described [32]. For the FcγR binding assay, 1μg/ml of antigen diluted in phosphate buffered saline (PBS) was coated onto 96-well flat bottom ELISA plates, followed by blocking with 1% BSA in PBS. The serum samples were tested in duplicate at 1/500 dilution, followed by the addition of biotinylated FcγRIIa or FcγRIII ectodomain at 0.2 μg/ml and 0.1 μg/ml respectively (all prepared in 1% BSA). FcγR binding was detected using streptavidin conjugated to horseradish peroxidase (HRP) at 1/10000 dilution in 1% BSA. Wells were incubated with 3,3’,5,5’-tetramethyl-benzidine (TMB) substrate, followed by 1M sulphuric acid, and absorbance was measured at an optical density (OD) of 450 nm. The FcγRIII probe represents binding activity of FcγRIIIa and FcγRIIIb.

Rabbit polyclonal IgG raised against CSP [31] was used as positive control to standardize for plate-to-plate variability, no serum control was used to determine background reactivity, and sera from malaria-naïve Melbourne adult blood donors were used as a negative controls (collected after informed consent). Positive responses were defined as reactivity greater than the mean plus 3×standard deviation of the Melbourne control samples. We also calculated the FcγR binding efficiencies relative to total IgG as follows: OD _FcγR binding_/OD _IgG_.

IgG, IgM and IgG subclasses were quantified by ELISA using standard methods in a previous study [17] and data were re-analysed in this study.

### Isolation of effector cells from peripheral blood for opsonic phagocytosis and ADCC

Isolation of whole leukocytes, neutrophils and peripheral blood mononuclear cells (PBMC) from peripheral whole blood were conducted using established methods [23].

Whole leukocytes were isolated by removing erythrocytes from peripheral blood using Dextran segmentation and hypotonic lysis approaches. Purified leukocytes were resuspended in RMPI-1640 supplemented with 10% fetal bovine serum and 2.5% heat inactivated malaria naïve human serum.

For PBMC and neutrophil isolation, peripheral whole blood was first separated by gradient centrifugation using Ficoll Paque (GE Healthcare). PBMCs were collected from the Buffy layer followed by 3 washes with cold PBS supplemented with 1% newborn calf serum. Neutrophils were isolated from the pellet by dextran segmentation enrichment and hypotonic lysis. The purified PBMCs and neutrophils were resuspended in RPMI-1640 supplemented with 10% fetal bovine serum and 2.5% heat inactivated malaria naïve human serum, respectively.

### Opsonic phagocytosis by neutrophils

The fluorescent latex beads (Sigma-Aldrich) were coated with full-length CSP using a published method [23]. Briefly, fluorescent beads with amine group modification on the surface were first incubated with glutaraldehyde to serve as a linker, followed by further co-incubation with CSP at concentration of 1mg/ml for antigen immobilization. The antigen coated fluorescent beads were opsonized with test sera at 1/100 dilution and then co-incubated with neutrophils for opsonic phagocytosis for 20 minutes at 37°C. Neutrophils were analysed using a BD FACS Canto II flow cytometer (gating strategy of flow cytometry data is reported in [23]). The level of phagocytosis was expressed as phagocytosis index, which was defined as the percentage of neutrophils that have ingested fluorescent beads. The phagocytosis index was further standardized as relative phagocytosis index as the percentage of the positive control, which was a rabbit polyclonal IgG raised against CSP [23]. Serum samples from non-malaria-exposed donors resident in Australia were used as controls. We have previously established the use of antigen-coated beads, in place of whole sporozoites, as a suitable model of opsonic phagocytosis [23].

### Opsonic phagocytosis in whole leukocyte assays, with primary monocytes or THP-1 cells

RTS,S induced opsonic phagocytosis by neutrophils and monocytes was compared using an established whole leukocyte assay [23]. Briefly, CSP-coated florescent beads were opsonized by a pool of sera selected from the RTS,S vaccination group at variable concentrations prior to co-incubation with whole leukocytes (isolated using the method above). Co-incubation was conducted at 37°C with 5% CO_2_. The plates were centrifuged at 4°C at the end of co-incubation and neutrophils were stained with anti-CD66b-AF647 (BD Bioscience) and monocytes were stained with anti-CD14-APC-H7 (BD Bioscience) and anti-CD16-BV421 (BD Bioscience) followed by quantification of phagocytosis using a BD FACS Verse flow cytometer. The beads phagocytosed by neutrophils were defined as PE^+^ and CD66b^+^ cells; the beads phagocytosed by monocytes were defined as PE^+^, CD14^+^ cells. The number of beads phagocytosed by neutrophils and monocytes were quantified based on their fluorescent intensity and standardized according to the total number of phagocytes (neutrophils and monocytes) that were analysed for each sample.

For quantification of opsonic phagocytosis by monocytes and THP-1 cells, CSP-coated fluorescent beads were opsonized with individual serum samples from the RTS,S vaccine group at concentration of 1/100 prior to co-incubation with THP-1 cells from culture or isolated PBMCs (contains monocytes) for 20 minutes. Monocytes were subsequently stained with anti-CD14-APC-H7 and the level of opsonic phagocytosis was analysed using a BD FACS Canto II flow cytometer and determined as the percentage of CD14^+^ cells that have phagocytosed beads (PE^+^), and the relative phagocytosis index was calculated as the percentage relative to the positive control (rabbit anti-CSP IgG) in the same plate. Assays using THP-1 cells have been previously reported [23] [33]. Standard THP-1 phagocytosis assays were performed using established methods that include 10% fetal calf serum, and additional assays including 2.5% human serum were conducted. The gating strategy of flow cytometry data for all cell types is reported in detail in a previous publication [23].

### Antibody-dependent cell-mediated cytotoxicity assay

The ADCC assay was performed using a previously published method [23]. Briefly, PBMCs containing NK cells were co-cultured with interleukin-2 overnight. CSP-coated beads were opsonized with a pool of sera selected from the RTS,S vaccination group. The opsonized beads were co-cultured with the primed PBMC and anti-CD107a-AF647 for 1 hour followed by addition of brefeldin A (Sigma-Aldrich) and protein transport inhibitor (BD Bioscience) and further co-cultured for 3 hours. After co-culture, the cells were stained with anti-CD3-APC-H7 (SK7, BD Bioscience), anti-CD56-PE-Cy7 (B159, BD Bioscience) and anti-CD16-BV421 (3G8, BD Bioscience). NK cells were defined as CD3^-^, CD56^+^, CD16+and the level of ADCC was quantified as the percentage of NK cells that were positive for CD107a staining by flow cytometry (LSR Fortessa X-20, BD Bioscience; gating strategy is shown in a previous publication [23]).

### Antibody dependent respiratory burst (ADRB) by neutrophils

Neutrophils were isolated from freshly collected venous blood using the EasySep Direct Human Neutrophil Isolation Kit (STEMCELL) according to manufacturer’s instructions, and used on the same day as collection. Neutrophil purity was assessed by cell morphology using light microscopy of Giemsa-stained slides of the isolated cells, viability of isolated cells was assessed using trypan blue exclusion.

CSP was coated on 96 well, white, flat bottom plates (NUNC) at 4 μg/mL in PBS and left overnight wrapped in parafilm in a damp box at 4°C. The liquid was then removed, and plates were washed with PBS followed by blocking with PBS 0.1% BSA (Sigma-Aldrich) for 1 hour at room temperature. Test serum samples (1/25 dilution in PBS), non-malaria exposed Melbourne controls (1/25 dilution in PBS), no sera (25 μl of PBS), or positive control polyclonal rabbit anti CSP antibody (1/500 dilution in PBS), was then added to each well and left at room temperature for 1-2 hours, plates were then washed 2 times with PBS. Just before addition to the plate, isolated neutrophils were resuspended in Hanks buffered salt solution at 2×10^6^ /mL and 20 μl of cells were added to each well followed by 20 μl of PBS with 33 ng/mL of HRP and 4 mM luminol (all from Sigma). The plate was centrifuged to settle the contents and then read immediately on a FLUOstar plate reader. Luminescence was measured in each well for 1 second every 2 minutes for 1 hour and was calculated as the average luminescence 5 minutes either side of the peak of the curve. Each plate was standardized using the rabbit anti-CSP and PBS controls which were included in duplicate on each plate. Relative luminescence was calculated for each sample as percentage relative to the positive control of the same plate.

### Statistics

All data were entered into Excel spreadsheets and subsequently imported into Stata SE 13.1 for statistical analysis. RTS,S induced responses (M0 versus M3) were compared by Wilcoxon paired rank sum test, Wilcoxon rank sum test for unpaired data, and Kruskal-Wallis Tests for antibody parameters between more than 2 groups. Spearman’s correlation was used to analyse the correlation between antibody parameters. Other specific tests were used as appropriate. For all analyses, the p values and statistical tests used are reported in figure legends and in tables.

Given the non-normal distribution of the data and presence of zero-values, generalised linear mixed modelling (GLMM; poisson distribution and log link function) was used to estimate latent growth-curve models exploring the subject-specific nature of the association between time and the magnitude and functions of antibodies. Generalised latent growth-curve models were estimated for each IgG subclass and functional antibody parameter using linear splines (based on the study measurement intervals); this was used to model the functional form of the effect of time (per month). The specific number of splines fitted for each model was based on visual interpretation of the functional form. Latent growth-curve models comprised two-levels, individuals at level-2 (i.e. random intercept and coefficient for linear spline limited to the 3-month period given heterogeneity observed in induction and estimation/model convergence/performance and parsimony considerations) and their levels and functions of antibody responses across-time at level-1. Post-estimation non-linear equations were used to estimate outcome half-lives and provide 95% confidence intervals about these estimates. Antibody induction and decay rates were estimated for each parameter specific time-interval applied in the modelling. The GLMM produced rate ratios (RR) and adjusted rate ratios (ARR) which represent the percent change in antibody magnitude or functional activity for each month increase in time for respective splines; i.e. RR <1 indicates a decrease (1 – RR ×100 gives % decrease) and >1 indicates an increase (RR – 1 × 100 gives % increase).

In order to estimate the longitudinal associations between the magnitude and functions of antibodies, latent growth-curve models were extended to include covariates in addition to linear splines (modelling the functional form of the effect of time), with unadjusted and adjusted effects estimated. RR and ARR were estimated and represent the percent change in the magnitude of an antibody parameter outcome for each unit increase in covariate parameter (E.g. percent change in FcγRIIa binding for each unit increase in IgG1). Model likelihood based information theoretic-criteria (Akaike Information Criteria [AIC] and Bayesian Information Criteria [BIC]) were used to determine model fit improvement when estimating time as a random effect (i.e. random slope for splines). To also assess the fit of the estimated latent growth-curve models, diagnostic plots comparing participants’ observed antibody levels with Bayesian model based (Best Linear Unbiased Predictions) predicted levels over time were produced and examined. In all statistical modelling, probability values were based on Wald statistics and the Huber/White sandwich variance estimator (i.e. robust standard error) was specified given the conditional variance was unlikely to equal the mean expectation in these data. Statistical inference was assessed at the 5% level. Stata version 15.1 statistical package (StataCorp LP, College Station, TX) was used in all statistical modelling.

## RESULTS

### RTS,S vaccine-induced antibodies interact with FcγRIIa and FcγRIII

Children received three RTS,S vaccine doses (at months 0, 1, and 2 of the study) and we initially compared antibodies in samples collected at month 3 (M3) versus pre-vaccination samples collected at baseline (M0). Children in the RTS,S group had significantly increased antibody-mediated FcγRIIa-binding and FcγRIII (IIIa and IIIb)-binding activity targeting CSP after vaccination (M3) compared to baseline (M0), or the comparator non-malaria vaccine group, in both the Manhiça and Ilha Josina study sites (p<0.001 for all tests; Figures 1A and 1B; Additional file Figures S1A and S1B). At baseline (M0), only few children had acquired antibodies to CSP, which were generally low in magnitude. Of note, the magnitude of FcγR-binding activity varied widely among children, and some children developed little or no IgG with FcγR-binding activity.

**Figure 1.**
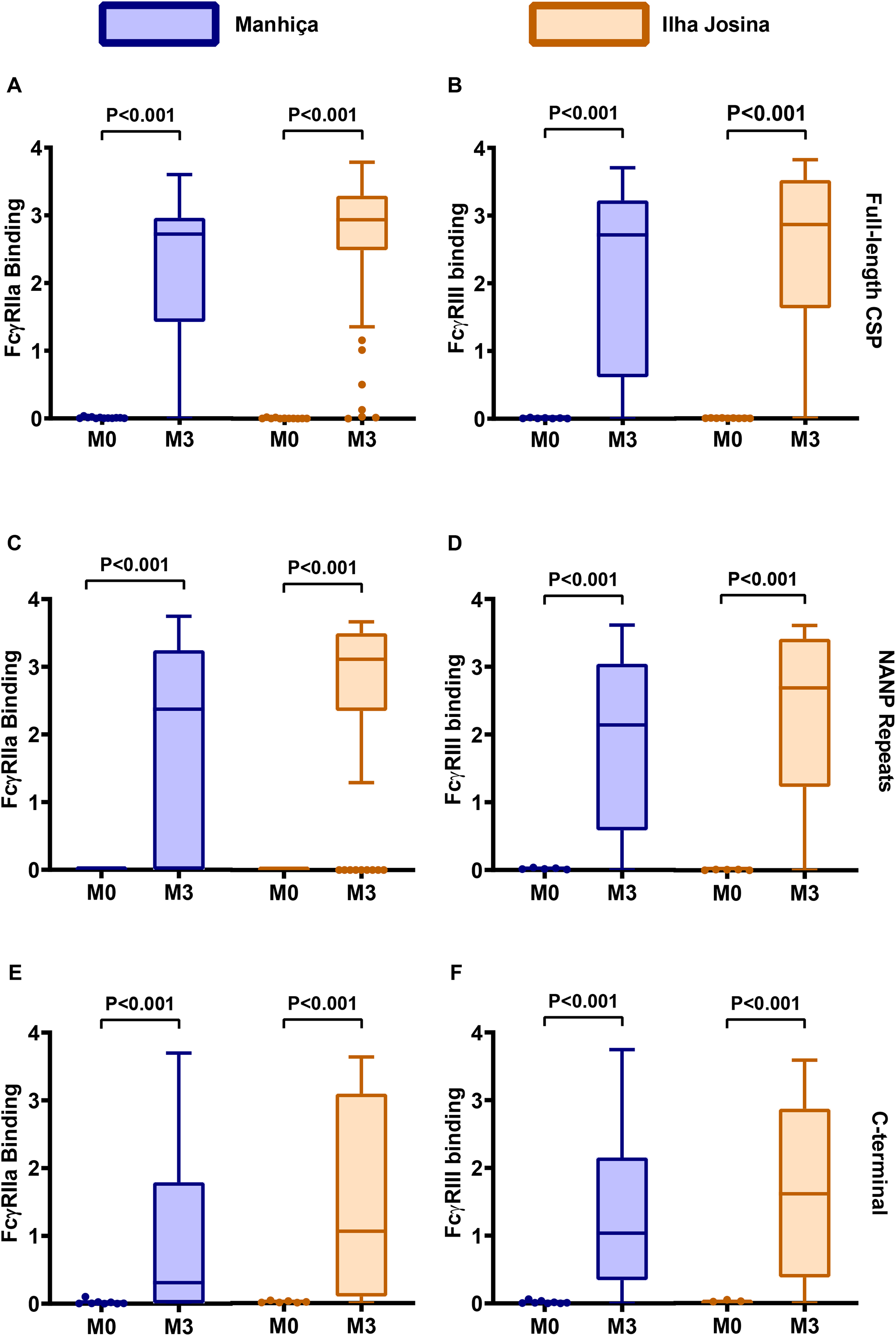
RTS,S vaccine-induced antibodies interact with FcγRs. Serum samples from children in the RTS,S vaccine group from Manhiça (blue box plots, n=50) and Ilha Josina (orange box plots, n=49) study sites were tested for antibodies that bind FcγRIIa (left panels) and FcγRIII (right panels). Samples collected at baseline (month 0, M0) and after the third vaccination (month 3, M3) were tested against (**A-B**) full-length CSP, (**C-D**) central-repeat (NANP) and (**E-F**) C-terminal (CT) regions of CSP. Boxes represent the interquartile range (IQR) with median (bar), and whiskers represent the highest and lowest values within 1.5× IQR. Values in the Y-axis indicate the optical density (OD) at 450nm. The percentage of children with a positive response are shown in Additional file Figure S1, and reactivities between paired samples were compared using the Wilcoxon’s signed rank sum test.

RTS,S-induced antibodies specific to the NANP and CT regions of CSP could promote binding of FcγRIIa and FcγRIII comparing M3 to M0, or compared to the comparator vaccine group (p<0.001 for all tests; Figures 1C-1F and Additional file Figures S1C-F). This observation was consistent in the Manhiça and Ilha Josina study sites. We evaluated the FcγRIIa and FcγRIII-binding activity relative to IgG for responses specific to each region of CSP, which was referred to as the FcγR-binding efficiency. RTS,S-induced IgG to NANP had a higher FcγRIIa and FcγRIII binding efficiency compared to IgG against the CT region (p<0.001, respectively, Additional file Figure S2).

### RTS,S vaccine-induced antibodies mediate cellular immune responses

The opsonic phagocytosis of sporozoites was recently found to be primarily mediated by neutrophils [23]. Evaluating a selection of children in the Manhiça cohort, we found that RTS,S vaccine-induced antibodies promoted neutrophil opsonic phagocytosis of CSP-coated beads (an established surrogate for sporozoites in phagocytosis assays [23]) (p<0.001 for M3 vs M0; Figure 2A). However, the magnitude of opsonic phagocytosis activity varied widely among children (median relative phagocytosis index (RPI) 35.08, interquartile range (IQR) 14.5-48.4%). Furthermore, these antibodies induced antibody-dependent respiratory burst by neutrophils, which indicates activation of neutrophils and the potential for cellular cytotoxicity [34] (p=0.011 for M3 vs M0; Figure 2B). RTS,S vaccine-induced antibodies also promoted opsonic phagocytosis of CSP-coated beads by the THP-1 monocyte cell line, which is commonly used as a model of monocyte phagocytosis *in vitro* (Figure 2C) [19, 20, 23, 33, 35, 36]. We confirmed that antibodies also effectively promoted phagocytosis with primary human monocytes purified from peripheral blood (Figure 2D). Previous studies reported that opsonic phagocytosis by THP-1 cells was largely mediated by FcγRI and that phagocytosis activity was partially inhibited by non-specific monomeric IgG present in human serum, which can bind to FcγRI on phagocytes [23]. To investigate this effect, we tested a pool of samples from RTS,S-vaccinated children for opsonic phagocytosis of CSP-coated beads by THP-1 cells in the presence and absence of non-immune human serum as a source of non-specific IgG. THP-1 cells could still promote substantial opsonic phagocytosis in the presence of non-immune human serum, although the magnitude was substantially reduced (Figure 2E). We further investigated the relative activity of neutrophils and monocytes in opsonic phagocytosis in a whole-leukocyte assay with fresh blood using a pool of samples from M3 versus M0 (Figure 2F). The rate of phagocytosis (number of beads phagocytosed per cell) was much higher by neutrophils than monocytes, which is similar to our previous observation of higher activity by neutrophils using naturally-acquired antibodies in adults and CSP-specific rabbit antibodies [23] Phagocytosis rate increased with increasing antibody concentrations for neutrophils and monocytes, and remained higher for neutrophils. Previously it has been reported that antibodies to CSP can promote NK cell activity through interacting with FcγRIIIa [18, 23]. Here, we confirmed that RTS,S-induced antibodies can induce NK cell activation characteristic of ADCC activity, comparing samples pooled from M3 versus M0 (Figure 2G).

**Figure 2.**
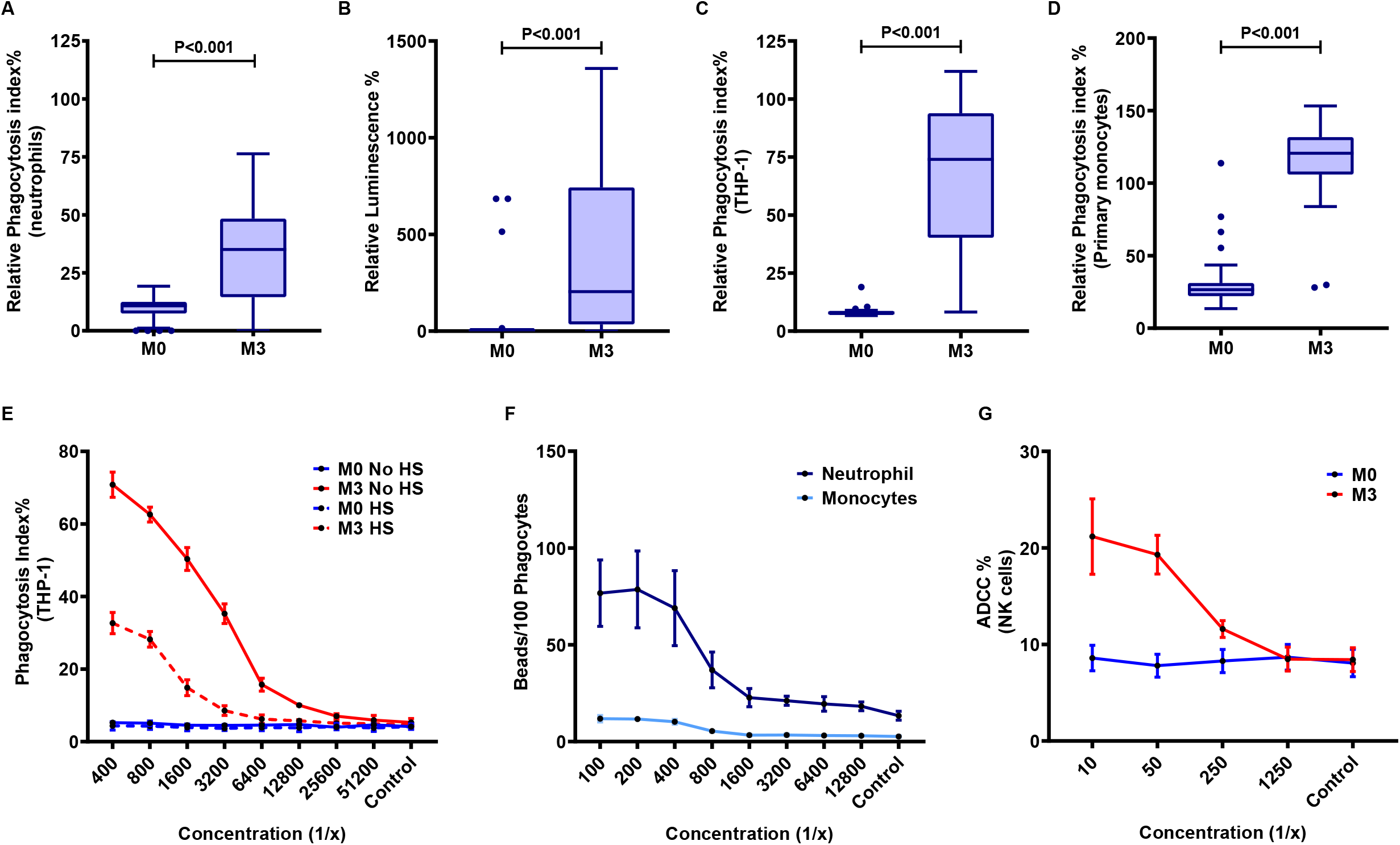
RTS,S vaccine-induced antibodies mediate cellular immune responses. (**A-C**) Serum samples from children in the RTS,S vaccine group (Manhiça cohort) collected at baseline (month 0, M0) and after vaccination (month 3, M3) were tested for the ability to promote (**A**) opsonic phagocytosis of CSP-coated beads by neutrophils (n=30), **(B)** antibody-dependent respiratory burst (OD units) by neutrophils against CSP (n=15), **(C)** opsonic phagocytosis of CSP-coated beads by THP1 cells (n=30). **(D)** Opsonic phagocytosis by primary monocytes (n=37 (22 from Manhica, 15 from Ilha Josina cohort)). Data are shown as box plots whereby boxes represent the interquartile range (IQR) with median (bar), and whiskers represent the highest and lowest values within 1.5× IQR. The reactivities between paired samples were compared using the Wilcoxon matched-pairs signed-rank test. Phagocytosis is reported as percentage relative to a positive control (rabbit IgG to CSP), ADRB is measured as luminescence units. (**E**) Pooled samples from M0 (blue lines or bars, n=99) and M3 (red lines or bars, n=99) were tested for opsonic phagocytosis of CSP-coated beads by THP-1 cells in the presence (dashed lines) and absence (solid lines) of non-immune human serum (HS). Dots and error bars represent the mean and standard error from two independent experiments. (**F**) Pooled samples from M0 and M3 (n=99) were tested for opsonic phagocytosis by neutrophils and monocytes in a whole leukocyte assay. Data show the number of CSP-coated beads phagocytosed by 100 cells. Dots and error bars represent the mean and standard error from two independent experiments using blood from two donors. Phagocytosis was higher for neutrophils than monocytes (P=0.003, two-way repeated measures ANOVA). (**G**) The same pooled samples from M0 and M3 were tested for activation of NK cells indicating ADCC activity. Y-axis indicates the percentage of NK cells that were positive for CD107a staining by flow cytometry (P<0.001, two-way repeated measures ANOVA). Dots and error bars represent the mean and standard error based on data from 4 experiments (different NK cell donors).

### The induction of FcγR-binding antibodies is associated with age and malaria exposure

The magnitude of RTS,S-induced FcγR-binding responses varied among children and we examined whether responses were influenced by age or malaria exposure, taking account of differences in malaria transmission intensity between the Manhiça and Ilha Josina study sites. Children were stratified into younger (12-24 months) and older (24-60 months) age groups (using the same age groups as were reported in the phase 2b clinical trial) [28]. In the lower malaria transmission site of Manhiça, no significant differences were seen in RTS,S-induced FcγRIIa and FcγRIII-binding activity between age groups (Figure 3A and 3B; Additional file Figure S3). However, in the higher transmission Ilha Josina study site, younger children had significantly higher FcγRIIa and FcγRIII-binding antibodies to CSP compared to older children (p<0.001; Figures 3A and 3B). Furthermore, FcγRIIa and FcγRIII-binding activity to NANP-repeat and CT regions was also significantly higher in younger children (p<0.001 for all tests; Additional file Figure S3). To assess whether the reduced induction of antibodies may be explained by higher malaria exposure level in the Ilha Josina cohort, we quantified IgG to blood-stage antigens AMA1 and MSP2, which are established biomarkers of malaria exposure [37]. We found that vaccine-induced FcγR-binding antibodies against CSP were lower among children with higher magnitude of IgG to AMA1 and MSP2 (Figure 3C, D; Additional file Figure S4). FcγR-binding antibodies were negatively correlated with IgG to MSP2 and AMA1 (FcγRIIa: Spearman’s correlation r=-0.356, p=0.011 for MSP2, r=-0.366, p=0.009 for AMA1; and FcγRIII: r=-0.362, p=0.010 for MSP2, and r=-0.356, p=0.011 for AMA1). We have previously reported that antibodies to AMA1 and MSP2 were higher in older children compared to younger children in the Ilha Josina study site [17]. This suggests that greater malaria exposure contributes to the reduced induction of FcγR antibodies by RTS,S vaccination.

**Figure 3.**
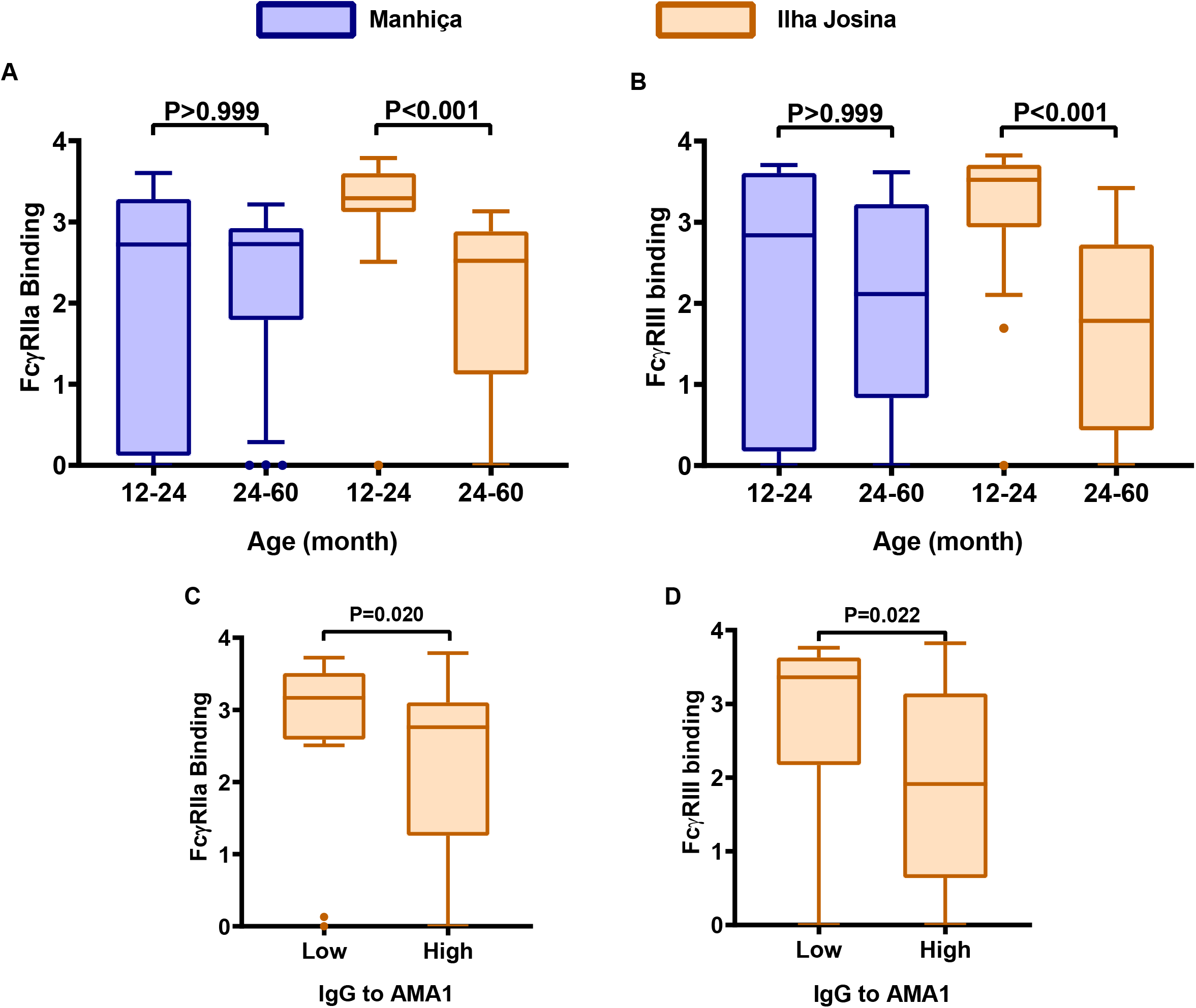
Induction of FcγR-binding antibodies to CSP in younger and older children. Serum samples from children in the RTS,S vaccine group were stratified into younger (12-24 months) and older (24-60 months) age groups from Manhiça (blue box plots; n=11 and n=39, respectively) and Ilha Josina (orange box plots; n=24 and n=26, respectively) study sites.Samples collected after vaccination at month 3 were tested for (**A**) FcγRIIa and (**B**) FcγRIII-binding to CSP. Boxes represent the interquartile range (IQR) with median (bar) and whiskers represent the highest and lowest values within 1.5× IQR. Y-axis data represent optical density (OD) at 450nm. The percentage of children with a positive response are shown in Additional file Figure S3, and reactivities between unpaired samples was compared using the Kruskal-Wallis rank sum test. For comparisons between Manhica and Ilha Josina: 12-24m age group, p=0.012 for FcγRIIa and p=0.091 for FcγRIII; for the 24-60m age group p>0.05 for both FcγRs. Results for the NANP-repeat and CT regions are shown in Additional file Figure S3. Serum samples from children in the Ilha Josina study site (n=49) were also tested for IgG to the merozoite antigen AMA1 which is an established biomarker of malaria exposure (**C, D**). Children were classified as having low or high AMA1 IgG based on being below or above the median. Children with low AMA1 IgG had significantly higher FcγR-binding antibodies; Mann-Whitney U-test.

### Relationships between different antibody responses

We examined relationships between different antibody parameters at M3 (Figure 4; Additional file Table S1). FcγRIIa and FcγRIII-binding to full-length CSP was significantly correlated with IgG to full-length CSP (Spearman’s correlation r=0.891 and 0.871, respectively; p<0.001; Figure 4A) and IgG to the NANP-repeat (r=0.794 and 0.775, respectively; p<0.001) and CT regions (r=0.590 and 0.658, respectively; p<0.001; Additional file Table S1). FcγRIIa and FcγRIII-binding to CSP also positively correlated with the corresponding FcγR-binding to the NANP repeat (r=0.547 and r=0.704, respectively; p<0.001; Additional file Table S1) and C-terminal regions (r=0.895 and r=0.958, p<0.001; Additional file Table S1). Furthermore, FcγRIIa-binding and FcγRIII-binding to full-length CSP were significantly correlated with each other (r=0.934, p<0.001); similar strength of correlations were seen in each study site (Figure 4B) and children’s age group (r=0.85-0.95; p<0.001; Figure 4B and C). Since antibody interactions with Fcγ-receptors can mediate cellular immune responses, we evaluated associations between FcγR-binding and neutrophil opsonic phagocytosis or ADRB. FcγRIIa-binding and FcγRIII-binding were each positively and significantly correlated with opsonic phagocytosis by neutrophils (r=0.603-0.610, p<0.001, Figure 4D) and ADRB (r=0.586-0.590, p<0.001; Figure 4E).

**Figure 4.**
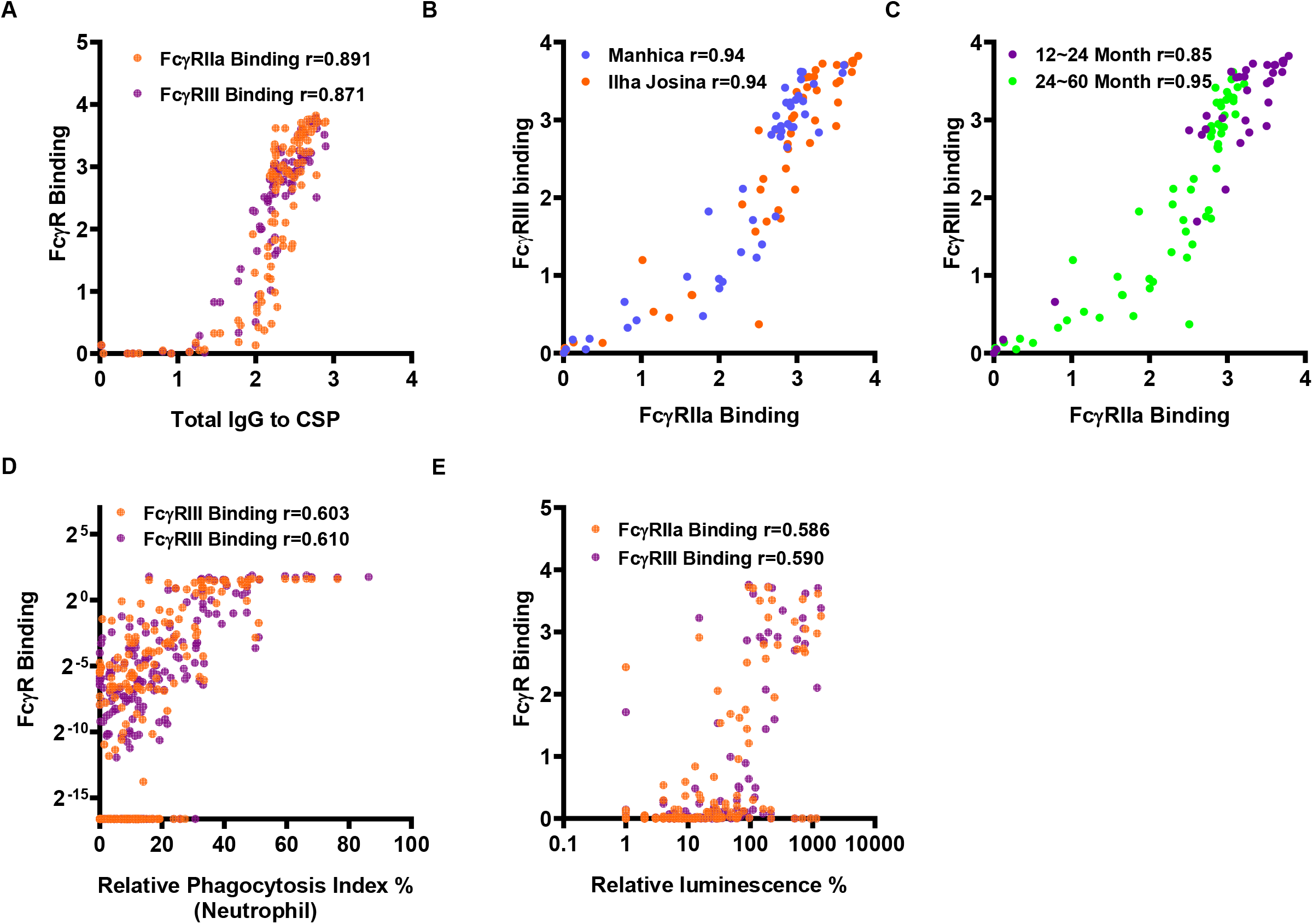
Correlations between antibody response types among children vaccinated with RTS,S. Serum samples from children vaccinated with RTS,S from the Manhiça and Ilha Josina study sites at M3 were tested for FcγRIIa/III-binding to CSP and serum samples from children in the Manhiça cohort were also tested for opsonic phagocytosis and ADRB. Individual samples are shown as the following scatter plots: (**A**) anti-CSP IgG and FcγRIIa or FcγRIII-binding (n=100); (**B**) FcγRIIa and FcγRIII binding by study site (Manhiça, n=50 and Ilha Josina, n=49) (**C**) FcγRIIa and FcγRIII binding to CSP by age group (younger, n=35 and older, n=65); (**D)** FcγRIIa or FcγRIII-binding and opsonic phagocytosis by neutrophils; (**E**) FcγRIIa or FcγRIII binding and ADRB. Correlations were evaluated using Spearman’s correlation coefficient (r). P-values <0.001 for all correlations. FcγR binding data are reported as optical density (OD) at 450nm.

### Differential decay of functional antibodies over time

Children from the Manhiça study site were sampled at multiple time-points for up to 5 years allowing the kinetics of RTS,S induced immunity to be assessed; these time points included months 3, 8.5, 21, 33, 45, and 63. All available data for each assay parameter and time-point were analysed using generalized linear mixed models (GLMM) to examine the rate of induction and decay for functional antibodies, anti-CSP IgG, IgG subclasses, and region-specific IgG (Figure 5, Table 1 and Additional file Figures S5 and S6 and Tables S2 and S3). FcγRIIa and FcγRIII-binding antibodies to CSP were strongly induced after RTS,S vaccination at M3, but substantially waned by M8.5 (approximately 6 months after vaccination) and were almost undetectable by M21 (Figure 5; Additional file Figure S5, S6). A similar trend was seen for ADRB activity by neutrophils. Although the induction of antibody-mediated neutrophils opsonic phagocytosis activity was modest, it showed a substantially slower decay, with responses persisting beyond M21 in some children. IgG subclass responses have been previously reported [17] and showed similar patterns of induction and decay. IgG to CSP, NANP and CT antigens generally showed a slower decay in comparison to functional antibody responses (Additional file Figure S6 and S7).

**Figure 5.**
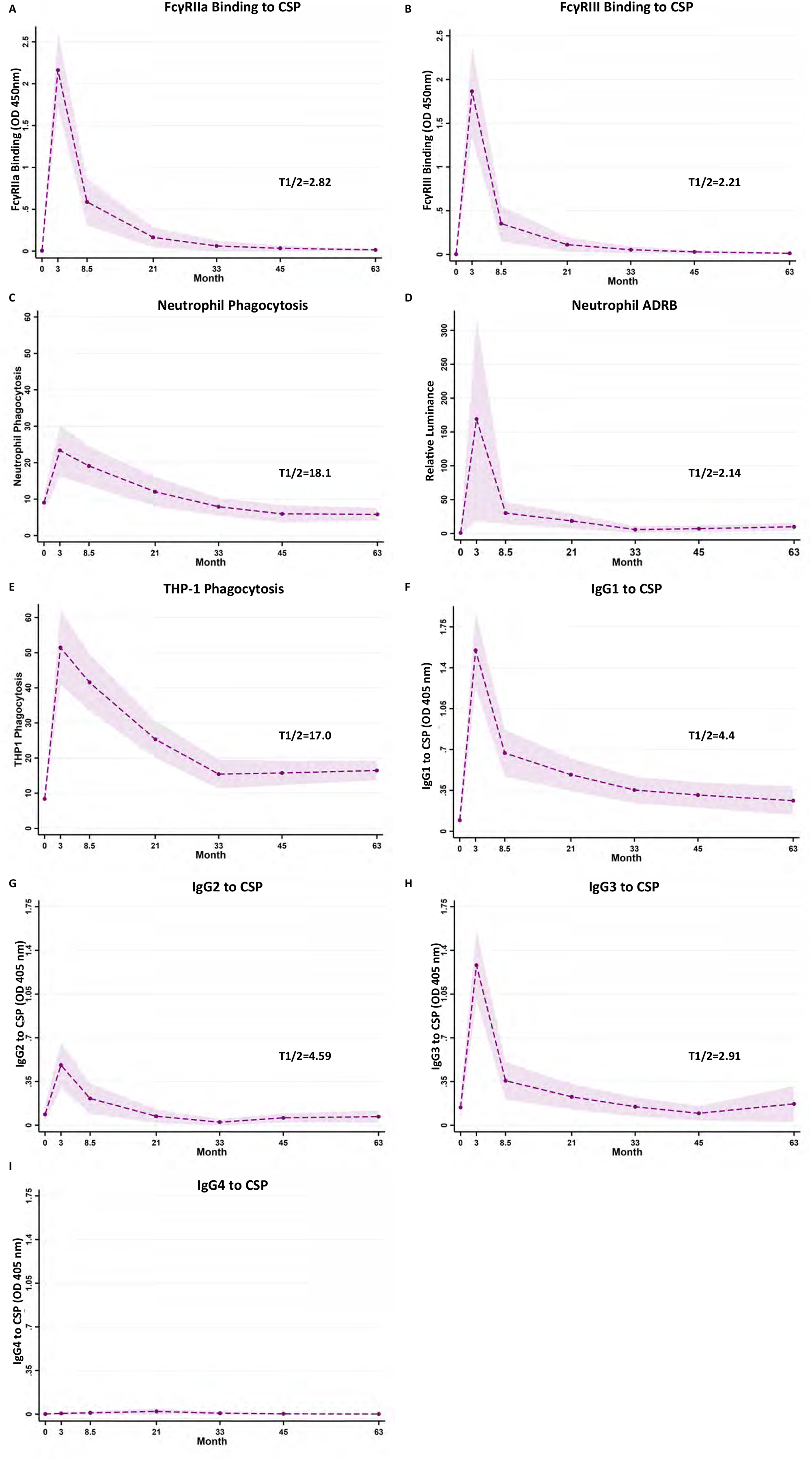
Kinetics of RTS,S vaccine-induced functional antibodies over time. A subset of children in the RTS,S vaccine group from the Manhiça study site were followed up over 5 years post vaccination. Serum samples were collected at baseline (month 0, M0), 30 days after the third vaccination (month 3, M3) and later time points (M8.5, M21, M33, M45 and M63). Samples were tested for **(A)** FcγRIIa-binding and **(B)** FcγRIII-binding to CSP, **(C)** opsonic phagocytis by neutrophils (n=33) and **(D)** antibody-dependent respiratory burst (ADRB) in neutrophils and (**E**) opsonic phagocytosis by THP-1 cells. Antibodies were also tested for IgG subclasses to CSP, including (**F**) IgG1, (**G**) IgG2, (**H**) IgG3 and (**I**) IgG4. Data from immunologic assays were analysed in generalized linear mixed model (GLMM). The dashed lines represent the predicted means, the shaded area represent the 95% CIs and T1/2 (in months) indicates the estimated half-lives from the generalized linear mixed model. Half-life represents the time for magnitude to reduce by 50% from the M3 time-point. Given the very low magnitude of IgG4 it was not possible to estimate a half-life for this parameter. Further details are provided in the Additional file, Table S1 and S2.

**Table 1.** Decay rate of RTS,S induced antibodies at different time points during follow-up.

| Antibody parameter | Follow-up time intervals |  |  |  |  |
| --- | --- | --- | --- | --- | --- |
|  | M3-M8.5 | M8.5-M21 | M21-M33 | M33-M45 | M45-M63 |
| FcγRIIa-binding | 19% | 10% | 8% | 5% | 5% |
| FcγRIII-binding | 25% | 9% | 6% | 5% | 5% |
| Neutrophil phagocytosis | 4% | 4% | 3% | 2% | *0% |
| Neutrophil ADRB | 28% | *4% | 9% | *-2% | *-2% |
| THP-1 cell phagocytosis | 4% | 4% | 4% | *0% | *0% |
| IgG1 | 11% | 3% | 3% | *1% | *1% |
| IgG2 | 13% | 9% | 9% | *-8% | *0% |
| IgG3 | 22% | 4% | 4% | 4% | *-4% |
| IgG4 | *-22% | *-6% | *9% | *8% | *5% |

Changes in RTS,S induced antibodies are reported as decay rate - percentage reduction in magnitude per month for each time interval.
See Table S2 in Additional file Materials for further details.
\*The p-value for all decay rates is <0.05, except where indicated with an asterisk

We initially used half-lives to summarise decay across antibody responses (representing the time taken for magnitude to reduce by 50% from M3; Figure 5, and Additional file Table S2). FcγRIII (*t*_*1/2*_*=* 2.21 months [95% CI=1.53-2.89]) and FcγRIIa (*t*_*1/2*_*=*2.82 months, [1.90-3.74]) binding and IgG3 (*t*_*1/2*_*=*2.91 months [2.13-3.69]) responses exhibited the shortest half-lives. IgG1 and IgG2 also had short half-lives (*t*_*1/2*_: IgG1 4.4 months, [2.9-5.9]; IgG2 4.5 months [1.1-8.1]). IgG4 was very low among children rendering it difficult to estimate *t*_*1/2*_. On the other hand, neutrophil phagocytosis had the longest *t*_*1/2*_ (18.1 months [17.8-18.36]) and *t*_*1/2*_ for THP-1 cells was 17.0 months (95%CI=13.5-20.5).

Decay of FcγR-binding activity and IgG subclasses was not uniform over the follow-up period and was generally greater from M3 to M8.5, and then more gradual and uniform over subsequent time (Table 1 and Additional file Table S3). For example, from M3-8.5, FcγR-binding decay showed a 22% and 27% reduction per month (for FcγRIIa and FcγRIII, respectively) and then more gradual after M8.5 (range 5 to 10% reduction per month). IgG1 and IgG3 reduced by 15% and 21% per month from M3-8.5, and subsequently by 0-4% per month after M8.5. Interestingly, the decay of neutrophil phagocytosis activity was more gradual and constant over the whole follow-up period after M3 (2 to 4% per month). This slower decay may reflect greater sensitivity for engaging immune complexes of FcγRs on the surface of neutrophils, which express FcγRIIa and IIIa/b [24]. There is also a background level of phagocytosis by neutrophils that occurs in the absence of specific antibodies [23]; therefore, neutrophil phagocytosis did not decay to zero as other antibody parameters did. Opsonic phagocytosis by THP-1 cells also showed a stable decay rate over follow-up (4% per month). During the follow-up period after completion of vaccination (M3), 23% (7/30) of children had malaria infection detected between M3-M8.5 and 30% (9/30) between M8.5-M21 time points. There was no significant difference in the magnitude of IgG, FcγRIIa or FcγRIII reactivity at M8.5 or M21 comparing those who did vs did not have malaria during follow-up. This is consistent with previously published data on RTS,S that suggests that infections do not boost vaccine antibodies [7, 9].

To further understand the decay of functional activity, we assessed FcγRIIa and FcγRIII binding efficiency of antibodies (FcγR binding activity relative to IgG magnitude) over time. RTS,S vaccine-induced antibodies were significantly less efficient in binding to FcγRIIa and FcγRIII at M8.5 months and later time-points (Figure 6A and 6B), compared to M3 (p<0.001). The decline in FcγR-binding efficiency was associated with a significant decline in the ratio of cytophilic IgG subclasses (IgG1 and IgG3, which promote FcγR-binding) to total IgG (p<0.001, Figure 6C and Additional file Figure S8), and with a decline in the ratio of cytophilic IgG subclasses to IgM (which does not bind FcγRs) (p=0.012, Figure 6D).

**Figure 6.**
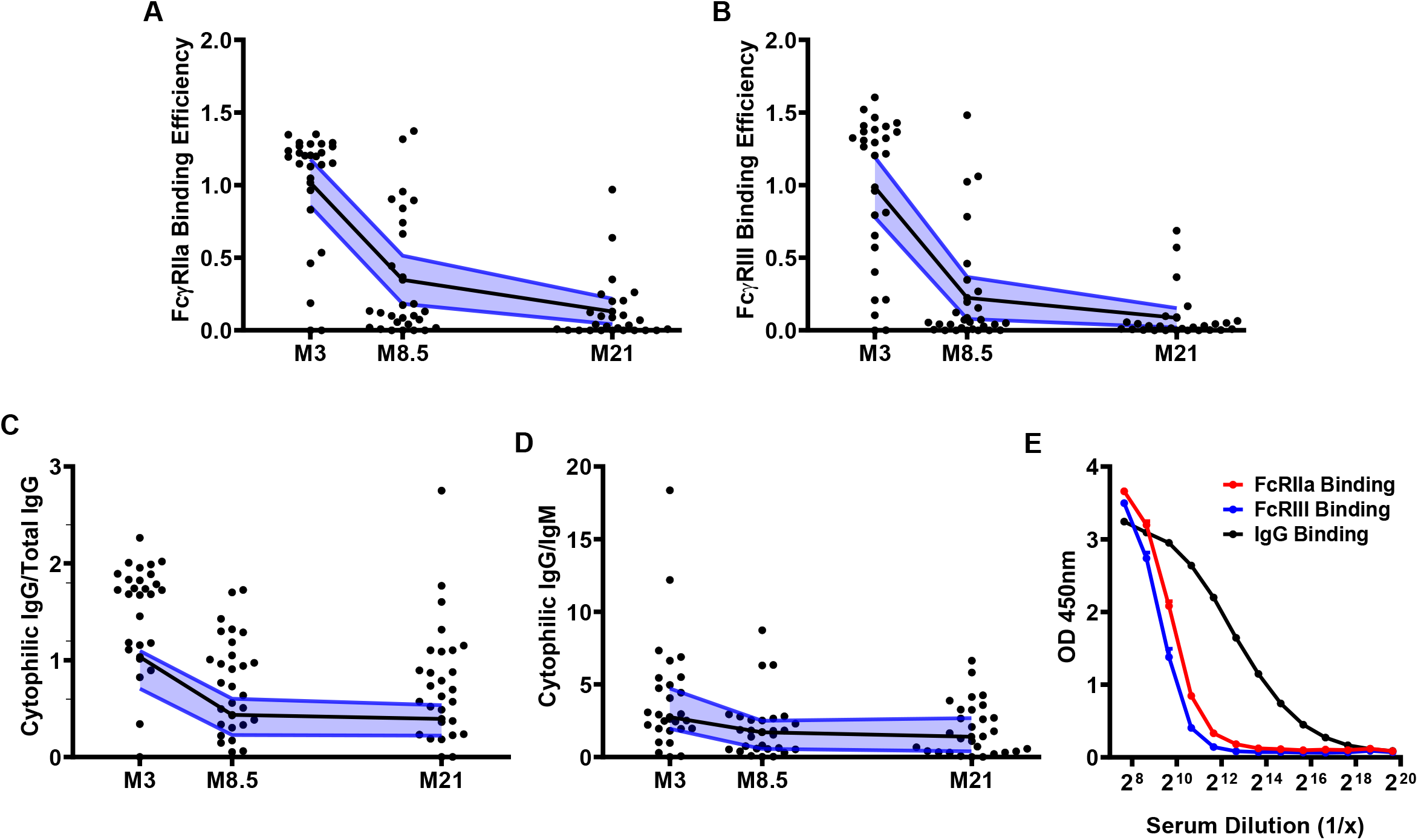
FcγR-binding efficiencies wane after RTS,S vaccination. Among children in the RTS,S vaccine group from the Manhiça study site, serum samples collected 30 days after vaccination (M3) and later time points (M8.5 and M21) were tested for various antibody responses to full-length CSP. FcγRIIa and FcγRIII binding efficiency was calculated as FcγR-binding relative to IgG magnitude for each sample at each time point. (**A**) FcγRIIa and (**B**) FcγRIII binding efficiency over time (n=28, Friedman test p<0.001, respectively); (**C**) Ratio of cytophilic IgG subclasses (IgG1 and IgG3) to total IgG (n=27, Friedman test p<0.001) and (**D**) Ratio of cytophilic IgG subclasses to IgM (n=26, Friedman test p=0.003). Later follow-up time points were not included because the magnitude of antibodies was very low. Individual samples are shown along with the median (black lines) and 95% CI (blue lines). (**E**) Pooled samples collected at M3 (n=99) tested at various dilutions for IgG (black line) and FcγR-binding to CSP (red line and blue line); dots and error bars represents mean and standard errors from 3 independent experiments.

Analyses of the relationship between IgG magnitude and FcγR binding also indicated there was a threshold effect of IgG binding before FcγR binding occurs. First, correlation plots between IgG and FcγR binding show that substantial FcγRIIa and FcγRIII binding only occurs above an IgG magnitude >1 OD units (Figure 4A); thereafter, the relationship appears to be largely linear. Second, we titrated a sample pool (from M3, n=99) in assays of FcγR binding versus IgG reactivity to CSP. FcγRIIa and FcγRIII binding dropped rapidly to baseline as IgG reactivity reduced and the slopes of the FcγR binding decay curves were greater than IgG (Figure 6E). These data suggest that as IgG declines over time, FcγRIIa and III-binding activity will be rapidly lost. The effect on neutrophil phagocytosis is less marked possibly because of the expression of multiple FcγRs on the neutrophil surface. Therefore, the decline of FcγR-binding efficiency of antibodies appears to be explained by a combination of a threshold effect and changes in the IgM and IgG subclass profile over time.

### Modelling the determinants of antibody functions over time

To understand and quantify the antibody types that determine functional activity, we extended our modelling approach, using antibody data from all time points over 5 years (M0 to M63) (summarised in Figure 7; details provided in Additional file Tables S4-S11). For FcγRIIa and FcγRIII binding activities, IgG1 and IgG3 exhibited the strongest associations among the IgG subclasses (Additional file Tables S4 and S5); rate ratios (RR) were estimated representing the percent change in participant FcγR-binding for a unit (OD) increase in IgG. For IgG1, a unit increase gave a 1.9-fold increase in FcγRIIa-binding and 2.3-fold increase in FcγRIII (p<0.001). For each unit increase in IgG3, we observed a 1.7-fold increase in FcγRIIa and 1.9-fold increase in FcγRIII binding (p<0.001). There were also significant associations observed for IgG2 with FcγRIIa and FcγRIII (RR=1.4 and 1.5, respectively; p<0.001). We extended the analysis by conditioning on each of the other IgG subclasses to explore independent associations between IgG subclasses and functional activity. In this analysis, IgG1 and IgG3 remained significantly associated with FcγRIIa and FcγRIII binding. Furthermore, IgG responses to both the NANP-repeat and CT regions were significantly associated with FcγR binding (FcγRIIa: RR=2.9 and 2.1 for NANP and CT respectively [p<0.001]; FcγRIII: RR=3.5 and 3.0 for NANP and CT respectively [p<0.001]) (Additional file Table S6 and S7). For neutrophil phagocytosis, IgG1 and IgG3 were significantly associated with activity, whereas associations for IgG2 and IgG4 were not significant (Figure 7; Additional file Table S8) (IgG1: RR=1.3, p<0.001; IgG3: RR=1.3, p=0.022). IgG to NANP (RR=1.5, p=0.039) and CT (RR=1.7, p<0.001) also exhibited associations with neutrophil phagocytosis (Additional file Table S9). Furthermore, antibody binding to FcγRIIa and FcγRIII showed significant associations with neutrophil phagocytosis (RR=1.3 and 1.4, respectively, p<0.001; Additional file Table S10), consistent with experimental *in vitro* data suggesting roles for these receptors in neutrophil phagocytosis of sporozoites [23]. Associations between THP-1 cell phagocytosis was significant for IgG1, IgG2, and IgG3 (RR=1.29, 1.23, and 1.29, respectively, Additional file Table S11); however, IgG2 was not significant in the adjusted analysis.

**Figure 7.**
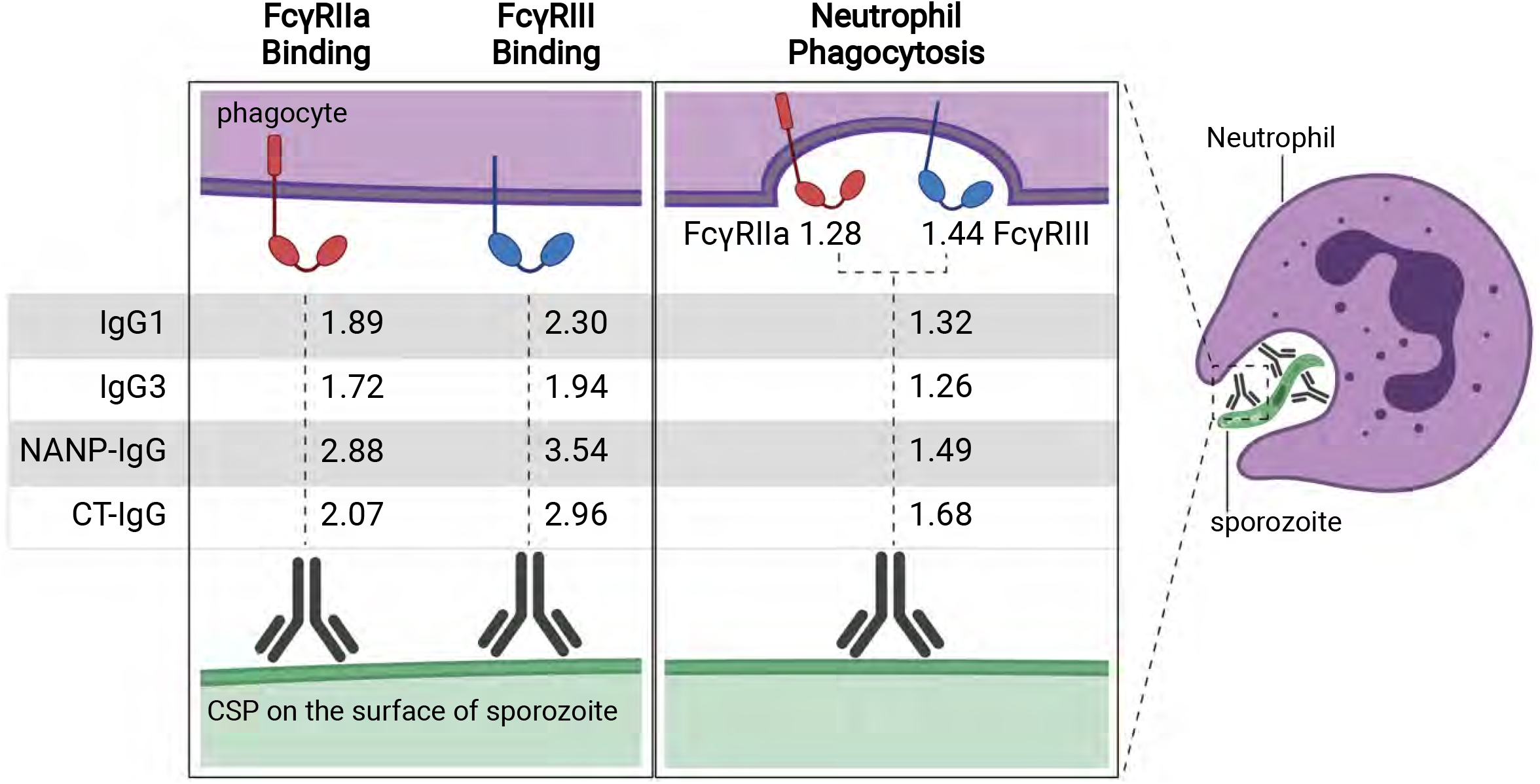
Antibody parameters correlated with functional activity. Antibodies against CSP opsonise sporozoites and interact with FcγRs to promote phagocytosis, which is predominantly mediated by neutrophils. Rate ratios are shown for the relationship between IgG subclasses and IgG to NANP-repeat and C-terminal domains of CSP, and antibody functional activities (FcγRIIa or III-binding and neutrophil phagocytosis). Rate ratios are also shown for the relationship between FcγRIIa and FcγRIII binding and neutrophil phagocytosis. Values are rate ratios and represent the percent change in participants’ antibody functional activities for each unit increase in IgG reactivity or FcγR binding. p≤0.001 for all rate ratios except neutrophil phagocytosis with IgG3 (p=0.023) and IgG to NANP (p=0.023). See Additional file Tables S4 to S11 for details.

## DISCUSSION

Here, we evaluated the induction, decay, and determinants of IgG effector functions in young children vaccinated with RTS,S in a malaria endemic region in Africa. Our study had several major new findings: 1) RTS,S vaccination induced IgG with FcγRIIa and FcγRIII binding activity and targeted the NANP repeat and CT regions of CSP. 2) Vaccine-induced antibodies promoted cellular functions including phagocytosis by neutrophils, THP-1 monocytes, and primary monocytes, and activation by NK cells for ADCC. However, responses were heterogeneous among children, and the magnitude of neutrophil phagocytosis by antibodies was relatively modest, which may be related to the modest vaccine efficacy. 3) The induction of functional antibodies was reduced among older children with higher malaria exposure. 4) Functional antibodies largely declined within a year post-vaccination, reflecting the decline in vaccine efficacy over that time. However, neutrophil phagocytosis activity showed greater persistence over time. Interestingly, the FcγR binding efficiency of antibodies (relative to total anti-CSP IgG concentration) also declined over time reflected by changes in relative concentrations of IgG1 and IgG3 and an IgG threshold effect for activity. 5) Biostatistical modelling of the determinants of functional activities, using data at all time points over 5 years, suggested IgG1 and IgG3 contribute similarly in promoting FcγR binding and phagocytosis, and IgG targeting both NANP-repeat and CT regions were important.

Response magnitude was highly variable among children (aged 1-5 years) and an important observation was that higher malaria exposure among children was associated with significantly reduced functional antibody activity. Addressing this might be important for achieving higher efficacy when RTS,S is implemented in some settings. Reduced vaccine responses may result from the impact that repeated malaria exposure can have on B cells, CD4+ T cells, and innate immune cell phenotypes and functions [38], which influence antibody generation. On the other hand, greater malaria exposure may be expected to generate antibodies to CSP, boost responses, and increase antibody affinity maturation, which could impact on functional activity. Age-related changes in immune function may also influence vaccine responses. Further studies are needed to understand these mechanisms and how they can be addressed to maximize vaccine responses in young children. Co-administration of malaria prevention with vaccine implementation may improve vaccine responses and efficacy [39]. Host genetics may also influence vaccine responses, and HLA genotypes were associated with RTS,S vaccine efficacy against experimental infection in malaria-naïve adults [40].

The interaction of IgG with FcγRs to promote phagocytosis, particularly by neutrophils, and other cellular functions against sporozoites appears to be an important contributor to malaria immunity [18, 23, 27]. This is the first time that the induction of these functional activities has been reported for any malaria vaccine in children or in a malaria-endemic population. Further, ADRB has not previously been assessed with any sporozoite antigen, or RTS,S, and it was interesting that robust ADRB was observed in some individuals. A role for ADRB in sporozoite clearance is plausible as evidence suggests singlet oxygen can inhibit growth of blood stage parasites *in vitro* [41, 42]. Further, antibodies promoted activation of NK cells, which occurs via engagement of FcγRIIIa, which may also contribute to immunity against sporozoites [18, 23]. Our data, and other published data [23, 43] suggest that neutrophils are substantially more active in phagocytosis than monocytes and are much more abundant than monocytes in blood (45–70% and 2–10% of peripheral blood white blood cells, respectively). This suggests neutrophils may play a more prominent role in clearance of sporozoites in blood. The role of NK cells and how they are activated to potentially clear sporozoites in RTS,S mediated immunity remains unclear and is a topic that warrants further research. While we studied neutrophils, monocytes, and NK cells from human blood, it will be valuable to study functions of these cell types in other tissues, including macrophages in the skin and liver (Kupffer cells) which may contribute to clearance of sporozoites. In animal models, antibodies to CSP can immobilise sporozoites in the skin [44], which may make them more susceptible to clearance by dermal phagocytes. Additional antibody mechanisms that may contribute to protection include complement fixation and inhibition of sporozoite traversal and invasion of hepatocytes [45]. Studies have shown that RTS,S induces complement-fixing antibodies in children and adults [17, 27]. There are limited data on the induction of invasion-inhibitory antibodies in RTS,S vaccine trials, or their correlations with protection. However, monoclonal antibodies isolated from RTS,S-vaccinated adults did have invasion inhibitory activity, suggesting this may be part of the protective mechanism [46, 47].

IgG1 and IgG3 have the highest activity for FcγR-binding and phagocytosis and are the dominant subclasses induced by RTS,S in children [48]. Prior studies found that these subclass responses were associated with protection from malaria in a sub-study of the phase III trial [48]. IgG subclass response profiles could be influenced to achieve greater functional activity and vaccine efficacy in the future through the selection of adjuvants, vaccine dosing and timing, and other factors. IgM does not engage FcγRs, but it is also induced by RTS,S. It was not associated with protection in children [48] or malaria-naïve adults with experimental infection studies [27]

IgG binding of the low affinity receptors FcγRIIa and FcγRIII is known to be influenced by epitopes targeted. The NANP-repeat and CT regions of CSP were both targets of functional FcγR-binding IgG. IgG to NANP-repeats broadly correlates with RTS,S efficacy and has been widely evaluated [16, 49-51]. At high concentrations, antibodies to the NANP-repeats can also inhibit sporozoite motility and invasion of hepatocytes *in vitro*, but this activity has not been shown to predict vaccine-mediated protection [10, 52]. Antibodies to the CT region have been less studied, but a recent analysis found that IgG to the CT region was associated with protection in the phase III trial [16]. Our findings indicate that antibodies to the CT region contribute to FcγR-mediated functions and may be an important component of RTS,S induced immunity, and it was recently shown that IgG to the CT region can promote phagocytosis of sporozoites [23]. This is important because the CT region is polymorphic and analysis of RTS,S phase III trial suggested that polymorphisms reduce vaccine efficacy [53, 54]. Generating antibodies with broad reactivity to different variants may be explored as a future strategy to improve RTS,S efficacy.

A key limitation of RTS,S is its relative short duration of efficacy, which largely wanes within 12 months after completion of primary vaccination [5]. Consistent with this, we found that FcγR-binding activity was largely lost over 18 months after completion of vaccination, and decay was more rapid in the first 6 months. The waning FcγR-binding was matched by declining magnitude of IgG1 and IgG3; the decline of IgG3 was more rapid than IgG1 and this subclass has higher FcγR-binding activity than IgG1 [55]. The decline in FcγRIIa and FcγRIII binding activity also reflected a threshold effect of IgG concentration for functional activity such that below a certain IgG concentration FcγR binding was lost. Neutrophil phagocytosis activity also declined substantially, but was better maintained over time than specific FcγR binding activity. It appears that the sensitivity of neutrophil phagocytosis to FcγR engagement by IgG means that there is a lower IgG threshold concentration required for activity, which may partly be explained by the expression of three activating FcγRs (FcγRIIIa, IIIb, and IIa) on the neutrophil surface. Our data is consistent with prior analyses of IgG, which suggest that there is not substantial boosting of antibodies through malaria infections occurring during the follow-up period [7]. However, larger studies with analysis of the genotypes of infections during follow-up will be required to better determine any boosting that may occur.

Loss of vaccine efficacy over time is a challenge for many malaria vaccines in development [10], but currently there are exceptionally limited data on the temporal kinetics of functional antibodies induced by RTS,S or any other malaria vaccine. Therefore, the data presented here contribute to understanding the durability of vaccine efficacy. We previously found that complement-fixing activity by antibodies, a mechanism implicated in immunity to sporozoites [31], also declined rapidly after vaccination in the same subset of children [17]. Potential strategies for improving vaccine durability include use of specific adjuvants, vaccine dosing and frequency, and improved targeting of key epitopes for optimal induction of functional antibodies. Given the lower decay rate of neutrophil phagocytosis, and evidence suggesting an important role in immunity [18, 23, 27], maximizing the induction of neutrophil phagocytosis activity of antibodies may be a potential future strategy to improve vaccine efficacy. Strategies to achieve this include increasing IgG3 or the ratio of cytophilic to non-cytophilic antibodies, such as through the use of adjuvants; increasing induction of antibodies to key epitopes that mediate phagocytosis; adding additional epitopes or antigens that promote phagocytosis (for example, targets of antibodies that promote phagocytosis were identified in the N-terminal sequence of CSP that is not in the RTS,S vaccine construct [23]).

We developed biostatistical models using the whole kinetic curve for each antibody parameter (all time points over 5 years) to evaluate factors associated with antibody functions. Our analyses highlighted the contributions of IgG1 and IgG3 for functional activity over time. While IgG3 has greater intrinsic activity for FcγR binding, it decayed more rapidly over time than IgG1. Our model supported antibodies targeting both regions of CSP for IgG interactions with FcγRIII and FcγRIIa and for neutrophil phagocytosis. While IgG responses to NANP-repeats have been a focus of vaccine development, our data suggest that improving responses to the CT regions, in addition to NANP-repeats, may be important for maximizing functional activity and possibly efficacy since antibodies to the CT region can promote phagocytosis of sporozoites. This is supported by recent analysis of a phase I/IIa infection challenge trial in malaria-naïve adults [20]. Glycosylation of IgG can influence FcγR binding activity [55]. Knowledge of the importance of this for malaria vaccine mediated immunity is unknown but should be investigated in future studies.

## Conclusions

Achieving high vaccine efficacy and greater longevity of efficacy are key global goals outlined by WHO and funding partners. We have demonstrated new functional antibody activities induced by RTS,S, and the temporal kinetics of these responses, in young African children, who are the primary target group of the vaccine. Our results provide insights to understand the modest and time-limited efficacy of RTS,S in children. In future studies, analysis of these functional antibodies needs to be extended into the RTS,S/AS01 phase III trial, including evaluation of responses in different populations, and an analysis of correlations of protection. Findings from this study address key knowledge gaps around RTS,S immunity and may contribute to improving RTS,S efficacy and longevity in the future or the development of next generation vaccines.

## Data Availability

The datasets used in the current study are available from the corresponding author on reasonable request and pending approval from ethics and regulatory committees where relevant. The Additional File (containing additional data analysis figures and tables) is available from the corresponding author on request. Specific materials and reagents used in immunologic assays can be provided dependent on availability.

## LIST OF ABBREVIATIONS

ADRB: Antibody-dependent respiratory burst
ADCC: Antibody-dependent cellular cytotoxicity
CSP: circumsporozoite protein
CT: C-terminal domain
FcγR: Fcγ-receptor
GLMM: generalised linear mixed modelling
M: month
PBS: phosphate buffered saline
RR: rate ratio; ARR – adjusted rate ratio
WHO: World Health Organisation

## DECLARATIONS

### Ethics approval and consent to participate

Ethics approval for this study and the parent clinical trial was obtained from Alfred Health Human Research and Ethics Committee (protocol number 174/18); Mozambique National Health and Bioethics Committee; Hospital Clinic of Barcelona Ethics Committee; PATH Research Ethics Committee. Parents/guardians of participants provided written informed consent.

### Consent for publication

Not applicable

### Availability of data and materials

The datasets used in the current study are available from the corresponding author on reasonable request and pending approval from ethics and regulatory committees where relevant. Specific materials and reagents used in immunologic assays can be provided dependent on availability.

## Authors’ contributions

GF, LK, EHA, performed the experiments and descriptive data analysis. PAA, FJIF developed and performed the statistical modelling. DRD, BDW, PMH, and JGB generated recombinant proteins and provided reagents, SJR provided methodologies. JS was involved in the clinical trial. GF, LK, CD, and JGB designed the study and led the data interpretation with input from all authors. GF, LK and JGB wrote the manuscript, with input from all authors. All authors read and approved the final manuscript.

## ACKNOWLEDGEMENTS

We thank all the study participants and their parents/guardians, and the staff of CISM and ISGlobal, especially Pedro Alonso, Joe Campo, Caterina Guinovart, Eusebio Macete, Pedro Aide and Augusto Nhabomba who were involved in the clinical trials. John Aponte, Ruth Aguilar, Gemma Moncunill and Clarissa Valim contributed to the study design. We also thank Nana Aba Williams, Núria Díez, Itziar Ubillos Marta Vidal and Laura Puyol for coordination, management and sample and data selection to facilitate the study. We thank Gennova Biopharmaceuticals and PATH MVI for providing recombinant circumsporozoite protein. Samples for this study were collected in a previous study [NCT00197041] funded by GlaxoSmithKline S.A. GlaxoSmithKline Biologicals SA was provided the opportunity to review a draft of this manuscript for factual accuracy, but the authors are solely responsible for final content and interpretation.

## ADDITIONAL FILE

Figure S1: Comparator vaccine does not induce antibodies that interact with FcγRs

Figure S2. FcγR-binding efficiencies of antibodies to the NANP-repeat and C-terminal regions of CSP

Figure S3. Induction of FcγR-binding antibodies to the central-repeat and C-terminal regions of CSP in younger and older children

Figure S4. RTS,S vaccine-induced FcγR-binding antibodies are negatively correlated with malaria exposure

Figure S5. Kinetics of RTS,S vaccine-induced functional antibodies over time Figure S6. Kinetics of RTS,S vaccine-induced IgG magnitude over time

Figure S7. Kinetics of RTS,S vaccine-induced IgG to the NANP and CT regions over time Figure S8. Decay of IgG1 and IgG3 relative to total IgG

Table S1. Spearman’s correlation coefficients between immune parameters that relate to FcγRIIa and FcγRIII binding among samples tested at M3

Table S2: Estimated half-life for IgG and functional factors

Table S3: Associations between log IgG subclass, log functional antibodies and time

Table S4-5: Associations between FcγRIIa and FcγRIII with IgG subclass

Table S6-7: Associations between FcγRIIa, FcγRIII, and IgG to NANP and IgG to CT

Table S8-10: Associations between opsonic phagocytosis by neutrophils with IgG subclass, IgG to NANP and IgG to CT, FcγRIII and FcγRIIa binding.

Table S11: Associations between opsonic phagocytosis by THP-1 cells and IgG subclass.

